# Saved and lost: The Health impact of routine vaccination and COVID-19-related restrictions in Democratic People’s Republic of Korea (1980–2024), a modelling study

**DOI:** 10.1101/2025.10.28.25338937

**Authors:** Hanul Park, Austin Carter, Young June Choe, Sempungu Joshua Kirabo, Minjae Choi, InBae Sohn, Joon Hee Han, Yo Han Lee

## Abstract

**Objectives:** Democratic People’s Republic of Korea has historically achieved high routine vaccination coverage, but these programs were substantially restricted during the COVID-19 pandemic. We estimated the cumulative averted disease burden from routine vaccination (1980–2024) and the excess burden associated with coverage declines during the pandemic (2021-2024).

**Methods:** We conducted a modeling study using WHO–UNICEF coverage data to estimate vaccine-preventable deaths and disability-adjusted life years (DALYs) for 8 pathogens. Estimates were based on mathematical models from the Vaccine Impact Modelling Consortium (VIMC), a global network of over 20 institutions, and the Global Burden of Disease (GBD) study. We applied the year-of-vaccination (YoV) approach to evaluate program performance and the excess burden, and used the calendar-year (CY) approach to assess the impact of catch-up vaccination. Based on historical coverage, pathogen-specific mortality, and vaccine efficacy, we quantified deaths and DALYs averted. To assess COVID-19-related disruptions, we compared actual coverage with a counterfactual scenario of uninterrupted vaccination to estimate the excess burden.

**Findings:** Between 1980 and 2024, routine vaccination averted an estimated 219 071 deaths (95% credible interval [CrI] 199 896–239 734) and 9.3 million DALYs, with hepatitis B (44·37%) and measles (28·57%) contributing most. Pandemic-related disruptions in 2021–2024 resulted in 15 624 excess deaths (11 379-20 747) and 649,565 DALYs (525 172-787 409) under the YoV method, partly mitigated by catch-up.

**Conclusion:** Routine vaccination has saved lives and prevented disability. The reduction in vaccination coverage associated with COVID-19 restrictions resulted in an excess burden that catch-up efforts cannot fully address.

## Introduction

The World Health Organization (WHO) Immunization Agenda 2030 (IA2030) aims to ensure equitable vaccine access by 2030, leaving no one behind regardless of life stage or circumstances^1^. Since 1974, vaccination has prevented 154 million deaths, with each death prevented providing an average of 66 years of full health^2^. However, the COVID-19 pandemic disrupted routine vaccination services in many countries, and if its impact continues through 2030, it is estimated that an additional 967,635 deaths could occur across 112 countries^3^.

The WHO recommended maintaining routine vaccination alongside infection prevention measures during the COVID-19 pandemic^4^. Nevertheless, immunization services were widely disrupted in many countries. While most countries experienced moderate and temporary declines in vaccine coverage, Democratic People’s Republic of Korea (DPRK) faced an unprecedented disruption^5^. DPRK exhibited distinct characteristics compared with countries that also implemented border closures, such as Australia and China. The country was highly dependent on vaccine imports, cold-chain logistics, and technical assistance from international partners. As a result of this reliance, the prolonged border closures likely resulted in vaccine stockouts and substantial disruptions to routine vaccination services.^6^

DPRK had steadily expanded its routine vaccination program for vaccine-preventable diseases (VPDs) since the early 2000s, achieving 96–98% coverage for most antigens by 2019^5^. However, coverage for multiple antigens sharply dropped in 2023, with DTP1 coverage declining from 98% in 2020 to 41% in 2023, and MCV1 coverage falling from 99% in 2020 to 28% in 2023 due to vaccine shortages according to the WHO-UNICEF Estimates of National Immunization Coverage (WUENIC). Based on WUENIC applied to population denominators from the UN World Population Prospects (UNWPP), we estimated that between 2021 and 2023, approximately 602,539 children remained zero-dose, having missed DTP1 vaccination (58.7% of the target population), while 361,139 missed MCV1 (35.2%)^7^. Similar declines were observed across other antigens^5^. Compared to global and regional trends, DPRK’s vaccination decline was disproportionately severe and sustained. Although a large-scale catch-up campaign was implemented in 2024, its impact remains to be fully evaluated^8^.

Although the health impacts of vaccination disruptions have been previously reported, no research has yet systematically quantified the disease-specific burden from the recent sharp coverage decline in DPRK. While data limitations of DPRK pose challenges to estimating vaccine-attributable burden, such analysis remains crucial for designing strategies to sustain vaccination during future crises. While some uncertainty remains due to limited local information, models from the Global Burden of Disease (GBD) project and the Vaccine Impact Modelling Consortium (VIMC) provided the most robust and transparent estimates for DPRK among the currently available data, as they were based on validated parameters within well-established modeling frameworks^3^,^9^.

This study focused on vaccine-preventable diseases included in DPRK’s national vaccination schedule, tuberculosis (BCG), diphtheria, pertussis, tetanus (DTP), hepatitis B (HepB), *Haemophilus influenzae* type b (Hib3), measles (MCV1), and rubella (RCV1), selected based on data availability and global disease burden. We aimed to quantify both the cumulative averted deaths and disability-adjusted life years (DALYs) of routine vaccination in DPRK since 1980 and the excess disease burden associated with the decline in vaccination coverage during the COVID-19 pandemic, using modeling approaches informed by VIMC and GBD estimates.

## Methods

### Study design

We estimated vaccine-averted deaths and DALYs in DPRK from 1980 to 2024 and quantified excess burden due to declines in routine vaccination coverage during the COVID-19 pandemic using our own mathematical and statistical models, which were informed by the methodological approaches of the VIMC and the GBD study. The year-of-vaccination (YoV) method, which assigns the lifetime benefits of each dose to the year of administration^10^, was applied to estimate lifetime vaccine benefits over 1980– 2024 and the excess burden during 2021–2024. To evaluate the short-term mitigating effect of catch-up vaccination in 2024, we used the calendar-year (CY) approach, which attributes impact to the year in which health outcomes occur^10^, providing a temporal view of realized burden. Both approaches are widely used in global vaccine impact assessments to capture programmatic benefits and time-specific disease burden.

In the CY approach, estimates may reflect cumulative effects of past vaccination, which may overestimate the results if interpreted as only the short-term impact of COVID-19-related disruptions. Presenting both CY and YoV allows capturing the cumulative long-term impact of immunization from past to present, programmatic effects at the time of vaccination, and the short-term mitigating effect of catch-up vaccination. We estimated YoV and CY through modeling based on VIMC and GBD data, incorporating vaccine efficacy, case incident rate (CIR), and case fatality rate (CFR). Further details are provided in Appendix A, B and C.

To estimate the excess disease burden due to missed vaccination, we constructed three scenarios. Scenario 1 (No-disruption) assumed that coverage levels from 2020 would be sustained through 2023, based on past trends and projected using an autoregressive integrated moving average (ARIMA) model. Scenario 2 (Disruption) applied the reduced coverage reported by WUENIC for 2020–2023. Scenario 3 (Disruption + Catch-up) assumed that children who missed vaccinations in 2021–23 received catch-up doses in 2024. The differences between Scenario 1 and Scenarios 2 or 3 represent excess deaths and DALYs attributable to vaccination disruptions (Appendix D). The study was approved by Korea University’s Institutional Review Board (KUIRB-2025-0452-01).

### Input data

Vaccination coverage data were obtained from the WUENIC, published in July 2025, and included all vaccines except polio-related vaccines for the years 1980 to 2024. To estimate vaccine impact in DPRK, we used estimates of cause-specific disease burden from the GBD 2021 study^10^. For each pathogen, CIR and CFR were adjusted to reflect the epidemiological context of DPRK, using 10-year historical data on incidence and mortality. Vaccine efficacy estimates were obtained from previously published literature (Appendix A). For YoV estimation, we used pathogen-specific disease burden data for measles, rubella, hepatitis B, and Hib from the VIMC. These estimates were derived using dynamic or static transmission models developed by 18 modelling groups across 112 countries^10,11^. VIMC data is stratified by age, year of vaccination, and vaccination activity type, and are suitable for attribution using the YoV method^10^. For vaccines against four pathogens not modeled by VIMC (i.e., tuberculosis, diphtheria, tetanus, and pertussis), we estimated averted deaths and DALYs based on GBD data (cause-specific disease burden, 2021 Socio-Demographic Index [SDI], and 2019 pathogen-specific Healthcare Access and Quality [HAQ] indices). We also used UNWPP 2024, released in July 2024, to obtain national-level, single-age population estimates for DPRK. Detailed data sources and adjustment procedures were described in Appendix B and C.

### 1980–2024: Estimating deaths and DALYs averted by vaccination

For the year-of-vaccination (YoV) method, antigen-specific impact ratio estimates from the Vaccine Impact Modelling Consortium (VIMC) and the number of fully vaccinated persons (FVPs) were used to calculate annual impact ratios (i.e., impact per FVP). The number of FVPs was calculated using national population estimates and vaccine coverage data^10,11^. To estimate a representative DPRK - specific impact ratio, time-weighted averages were calculated, placing greater weight on recent years to reflect changes in epidemiology and vaccination programs. For vaccines not modeled by VIMC, we estimated averted deaths and DALYs using relative risks (RRs) derived from inverted vaccine efficacy and all-cause mortality. Annual, cause-specific RRs were estimated based on available GBD mortality data, and for years beyond data availability (i.e., post-2021), RRs were extrapolated using logistic regression models incorporating year, SDI, and HAQ. Uncertainty was quantified using 1,000 bootstrap iterations and 10,000 Monte Carlo simulations. For the CY method, cause-specific disease burden data from the GBD study were used to estimate hypothetical burdens in the absence of vaccination by applying vaccine efficacy and coverage levels. Averted deaths and DALYs were then calculated by comparing these counterfactual burdens with the observed GBD disease burden^10^. Detailed estimation procedures, including parameter values and formulas, are provided in Appendices B and C.

### 2021–2024: Estimating excess deaths and DALYs due to restricted routine vaccination during the COVID-19 pandemic

The YoV method was used to estimate the disease burden during the COVID period in the same methods as the 1980-2024 vaccine impact was calculated. To estimate excess burden for the period 2021–2024 under Scenario 2 (disruption), we first estimated the hypothetical number of deaths in the absence of vaccination. Using this, we calculated averted deaths and DALYs for Scenario 1 (no disruption) and Scenario 2. The difference between these scenarios was then used to quantify the excess burden attributable to coverage decline. For Scenario 3 (Disruption + Catch-up), we first estimated the averted burden under the disruption scenario and then added the averted burden due to catch-up vaccination. The catch-up averted burden was derived by estimating the catch-up target population—those who missed vaccination during 2021-2023, and adjusting for survival probabilities considering the CIR, CFR, and vaccine efficacy. Finally, the difference in averted burden between Scenario 1 and Scenario 3 was calculated to estimate the excess burden after catch-up efforts. Detailed calculation procedures are provided in the Appendix B and C.

## Results

Since the late 1980s, DPRK maintained vaccination coverage above 90%, with a brief decline during the 1995 economic crisis, then high coverage across most antigens after 2000 (Appendix D.1). YoV framework, which attributes all averted deaths to the year when vaccination occurred, routine vaccination targeting eight pathogens between 1980 and 2024 is estimated to have averted 219 071 deaths (95%CrI 199 896–239 734) and added 9 347 039 years (95%CrI 8 766 861–9 956 199) of full health, measured in DALYs (Table 1, Figure 1). Hepatitis B vaccination accounted for the largest number of lives saved, 97 241 deaths (95%CrI 83 311–112 177; 44·37%), followed by measles vaccination, which prevented 62 491 deaths (95%CrI 57 909–67 357; 28·57%). Regarding total years of full health gained, measles vaccination contributed the largest share (46·12%), reflecting the relatively high morbidity associated with measles despite its lower mortality (Table 1, Figure 1).

**Table 1.**
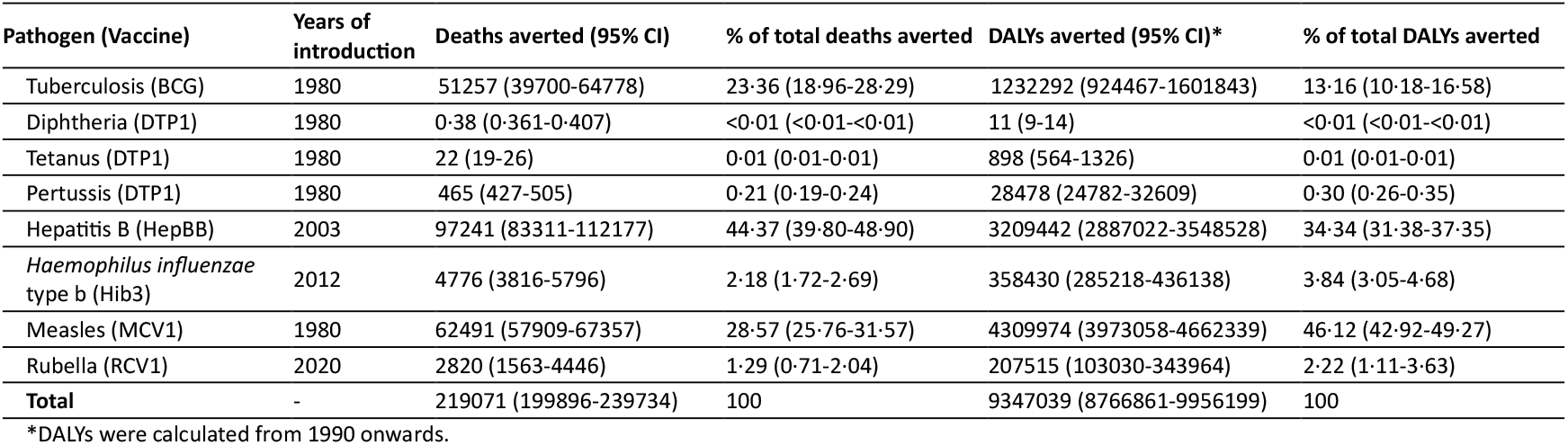
Estimated cumulative deaths and DALYs averted by routine immunization in DPRK, 1980–2024 (years of vaccination)

**Figure 1.**
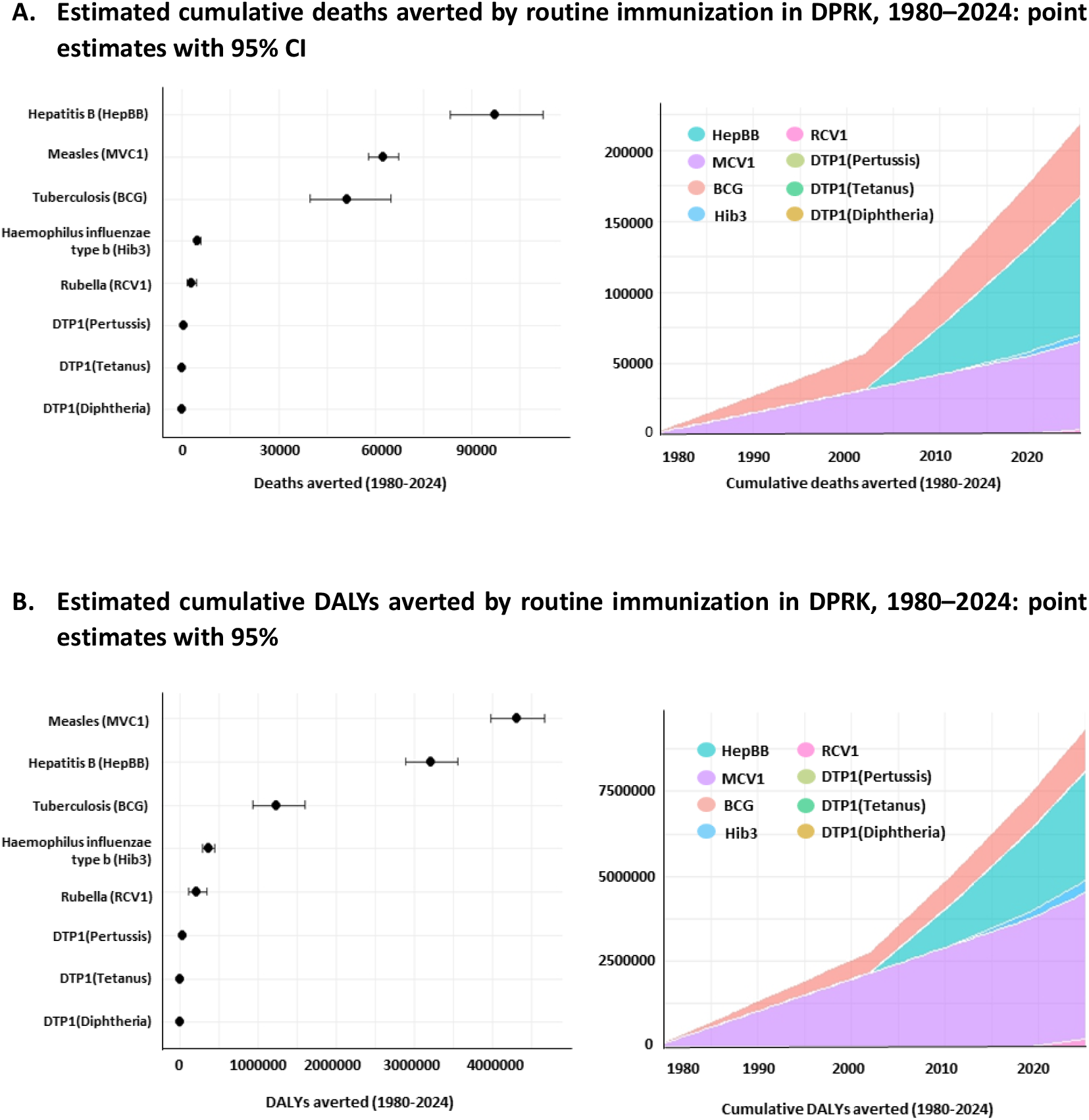
Estimated cumulative deaths and DALYs averted by routine immunization in DPRK, 1980–2024 (years of vaccination)

However, coverage declined sharply from 2020, particularly in 2021–2023, due to COVID-19-related disruptions (Appendix D.2). Between 2021 and 2024, these disruptions led to an estimated 15 624 excess deaths (95%CrI 11 379–20 747) and 649 565 DALYs (95%CrI 525 172–787 409) lost, representing 41·8% and 39·7%, respectively, of the deaths and DALYs that would have been averted in the absence of the collapse. Hepatitis B accounted for the largest share of excess 11 641 deaths (95%CrI 7 443–16 679), followed by measles with 1 972 (95%CrI 1 356–2 726). These correspond to 54·15% and 26·02% of the counterfactual averted deaths in the absence of vaccination disruption. Similarly, Hepatitis B contributed most to the DALYs lost (383 940 years, 95%CrI 285 724–496 513), with measles second (136 029 years, 95%CrI 90 070–193 860), (Table 2, Figure 2).

**Table 2.** Estimated cumulative excess deaths and DALYs from routine immunization restrictions due to the COVID-19 pandemic in DPRK, 2021–2024 (years of vaccination)

| Pathogen & Vaccine | 2021-2024 Scenario2: Disruption |  |
| --- | --- | --- |
|  | Excess deaths (95% CI) | Excess DALYs (95% CI) |
| Tuberculosis (BCG) | 470 (80-1458)<br>%: 12.03 (2.97-28.00) | 14852 (2514-46150)<br>%: 12.15 (2.90-28.26) |
| Diphtheria (DTP1) | <0.01 (<0.01-<0.01)<br>%: 44.78 (38.55-50.06) | <0.01 (<0.01-<0.01)<br>%: 45.04 (38.90-50.30) |
| Tetanus (DTP1) | 0.02(0.01-0.02)<br>%: 41.82 (31.34-50.64) | 0.7 (0.4-1.0)<br>%: 41.98 (31.53-50.69) |
| Pertussis (DTP1) | <0.01 (<0.01-<0.01)<br>%: 48.30 (43.75-52.10) | 0.04 (0.03-0.06)<br>%: 48.29 (43.81-51.98) |
| Hepatitis B (HepBB) | 11641 (7443-16679)<br>%: 54.15 (42.21-65.26) | 383940 (285724-496513)<br>%: 54.15 (45.71-62.10) |
| *Haemophilus influenzae* type b (Hib3) | 952 (587-1418)<br>%: 60.02 (45.53-72.83) | 71402 (44013-106295)<br>%: 60.01 (45.48-72.86) |
| Measles (MCV1) | 1972 (1356-2726)<br>%: 26.02 (19.46-33.25) | 136029 (90070-193860)<br>%: 26.03 (18.89-33.72) |
| Rubella (RCV1) | 590 (205-1213)<br>%: 26.11 (11.47-43.63) | 43341 (11740-95923)<br>%: 26.04 (9.32-46.03) |
| **Total** | 15624 (11379-20747)<br>%: 41.82 (31.34-50.64) | 649565 (525172-787409)<br>%: 39.68 (34.09-45.19) |

%: Proportion of excess burden relative to the averted burden in the non-disruption scenario

**Figure 2.**
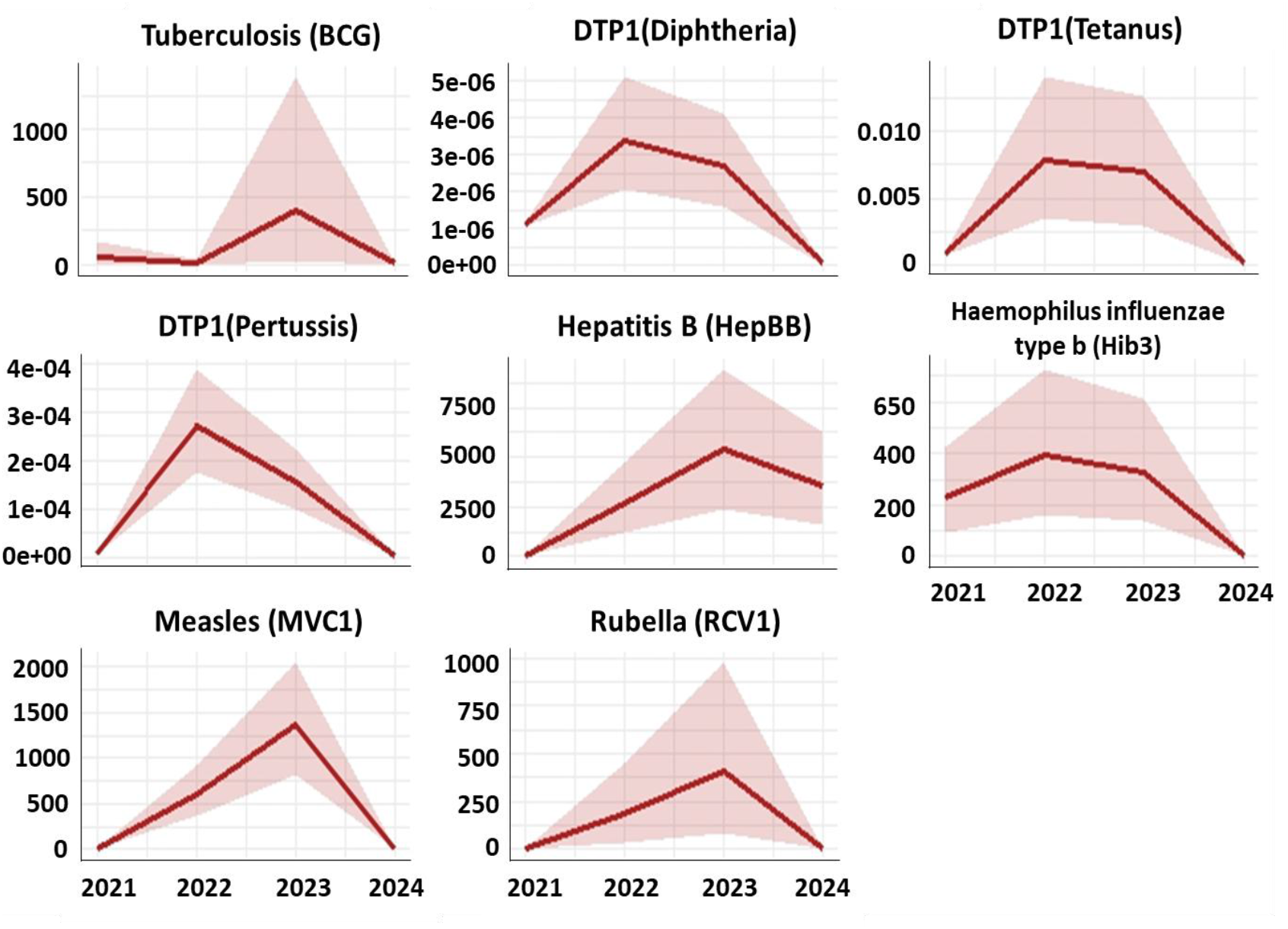
Estimated cumulative excess deaths from routine immunization restrictions due to the COVID-19 pandemic in DPRK, 2021–2024 (years of vaccination)

Under the CY framework, which attributes disease burden to the year in which deaths would have occurred, we compared Scenario 1 to Scenario 2 and Scenario 3. Between 2021 and 2024, the difference between Scenario 1 and Scenario 2 corresponds to 113 044 excess deaths (95%CrI 82 476–159 800) and 3 896 494 DALYs (95%CrI 2 845 203– 5 529 932), (Table 3). With catch-up vaccination, these figures would have been reduced slightly to 111 766 excess deaths (95%CrI 82 406–160 771) and 3 883 363 DALYs (95%CrI 2 851 493– 5 530 667), indicating the partial mitigation of health losses. In both scenarios, hepatitis B and tuberculosis accounted for the largest contributions to excess deaths and DALYs (Table 3, Figure 3).

**Table 3.** Estimated cumulative excess deaths and DALYs from routine immunization restrictions during the COVID-19 pandemic in DPRK, 2021–2024 (calendar year)

| Pathogen & Vaccine | 2021-2024 Senario2: Disruption |  | 2021-2024 Senario3: Disruption + Catch up |  |
| --- | --- | --- | --- | --- |
|  | Excess deaths (95% CI) | Excess DALYs (95% CI) | Excess deaths (95% CI) | Excess DALYs (95% CI) |
| Tuberculosis (BCG) | 11314 (3090-29368)<br>%: 10.97 (3.59-23.19) | 352327 (96982-928035)<br>%: 11.00 (3.76-23.43) | 10124 (3044-30124)<br>%: 11.04 (3.65-23.68) | 334118 (95831-962984)<br>%: 10.94 (3.80-23.55) |
| Diphtheria (DTP1) | 15 (12-21)<br>%: 45.66 (37.53-50.53) | 832 (634-1150)<br>%: 45.83 (37.67-50.65) | 15 (12-21)<br>%: 45.50 (37.51-50.39) | 832 (638-1149)<br>%: 45.92 (38.29-50.70) |
| Tetanus (DTP1) | 758 (280-1193)<br>%: 42.07 (22.97-55.42) | 31978 (11893-50019)<br>%: 41.77 (22.95-55.45) | 758 (277-1192)<br>%: 41.92 (22.65-55.31) | 31860 (11852-50018)<br>%: 42.04(23.15-55.38) |
| Pertussis (DTP1) | 32 (24-44)<br>%: 44.44 (37.63-49.40) | 3103 (2311-4265)<br>%: 44.11 (37.18-49.20) | 20 (13-32)<br>%: 27.96 (21.21-35.05) | 3093 (2303-4264)<br>%: 44.05(37.18-49.06) |
| Hepatitis B (HepBB) | 90589 (65370-133036)<br>%: 54.71 (43.62-64.63) | 2824238 (2047875-4113733)<br>%: 54.05 (43.30-63.75) | 90463 (65372-132829)<br>%: 54.78 (44.02-64.57) | 2827848 (2050977-4161651)<br>%: 54.08(43.10-63.83) |
| *Haemophilus influenzae* type b (Hib3) | 6159 (5048-7690)<br>%: 55.80 (45.03-63.70) | 350798 (285787-439661)<br>%: 56.27 (46.13-64.20) | 6147 (5029-7700)<br>%: 55.60 (45.20-63.65) | 351035 (286581-438714)<br>%: 56.32 (45.73,64.08) |
| Measles (MCV1) | 0.001(0.001-0.002)<br>%: 30.09 (23.86-36.87) | 0.003 (0.002-0.005)<br>%: 15.87 (10.72-21.92) | -0.003 (-0.018-0.001)<br>%: -81.47 (-498.64-35.94) | -0.001 (-0.016-0.004)<br>%: -3.71(-78.31-20.10) |
| Rubella (RCV1) | 4177 (771-13812)<br>%: 32.34 (5.98-62.23) | 333218 (62626-1022149)<br>%: 31.94 (6.13-62.55) | 4239 (771-13452)<br>%: 32.34 (5.92-62.87) | 334577 (62194-1057150)<br>%: 32.10 (5.99-62.72) |
| **Total** | 113044 (82476-159800)<br>%: 40.33 (27.68-50.31) | 3896494 (2845203-5529932)<br>%: 39.79 (27.67-49.65) | 111766 (82406-160771)<br>%: 40.33 (27.02-50.45) | 3883363 (2851493-5530667)<br>%: 39.85 (27.41-50.02) |

%: Proportion of excess burden relative to the averted burden in the non-disruption scenario

**Figure 3.**
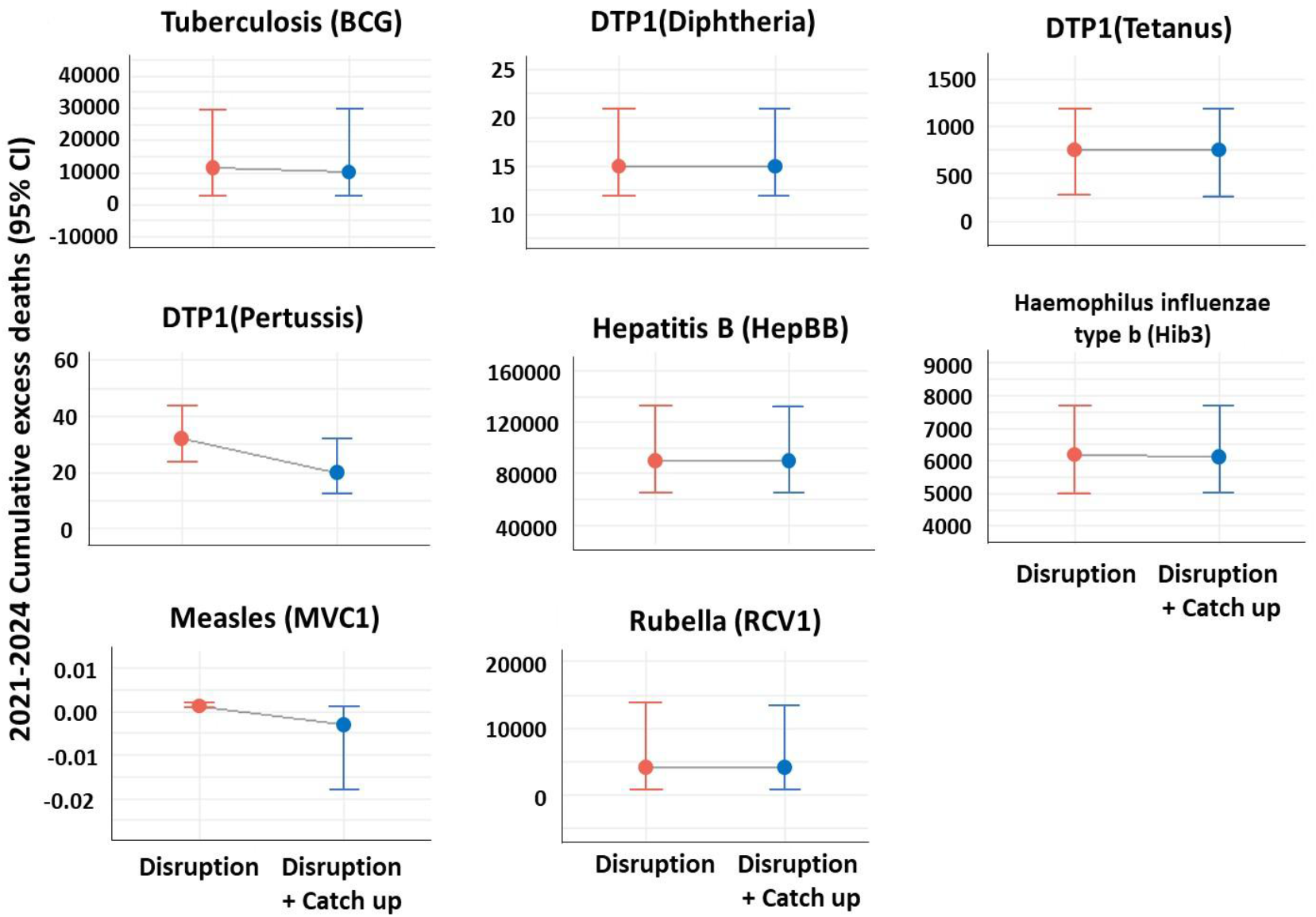
Estimated cumulative excess deaths from routine immunization restrictions during the COVID-19 pandemic in DPRK, 2021–2024 (calendar year)

## Discussion

This study provides a comprehensive assessment of the health impact of routine vaccination in DPRK over four decades. We estimate that between 1980 and 2024, routine vaccination averted approximately 219 071 deaths and preserved 9.3 million DALYs, with hepatitis B and measles contributing the largest shares of the total impact. However, the unprecedented disruptions during the COVID-19 pandemic between 2021 and 2023 likely resulted in 15 624 excess deaths and a loss of 649 565 healthy life years. In the CY approach, we estimated that catch-up vaccination in 2024 partially mitigated this excess burden. These findings indicate that immunization setbacks cause immediate excess deaths that catch-up efforts cannot fully reverse, highlighting the critical importance of ensuring that routine vaccination programs are protected and maintained even in the face of crises.

Among the 219 071 averted deaths, hepatitis B, measles, and tuberculosis accounted for the largest shares, with 97 241, 62 491, and 51 257 deaths averted, respectively. This averages approximately 4 868 deaths per year (1·9% of recent annual mortality) in DPRK. These estimates highlight the substantial impact of routine immunization, although factors such as vaccine timing and target ages should be taken into account. The cumulative averted deaths (1980–2024) observed in our study align with multi-country modelling. For example, VIMC estimated 97 million deaths averted across 112 low- and middle-income countries during 2000–2030, mainly from measles and hepatitis B^3^. Similarly, our results showed a dominance of hepatitis B and measles, with a notable contribution from tuberculosis, not included in the VIMC analysis. Despite differences in study period and pathogen coverage, our estimates provide a context-specific perspective on the long-term averted burden in DPRK and appear broadly plausible in light of comparable modelling evidence.

BCG vaccination, introduced in 1980, primarily protected against severe forms of childhood tuberculosis ^12^. In contrast, hepatitis B vaccination, introduced later in 2003, contributed the largest number of deaths averted, reflecting both the high endemicity and the long-term burden of chronic hepatitis B infection in North Korea, potentially including prevention of vertical (mother-to-child) transmission. Measles, as an acute, outbreak-prone infection with high transmissibility and childhood mortality risk, has been substantially mitigated through routine MCV since 1980, underscoring the importance of herd immunity. More recently introduced vaccines, such as Hib3 and RCV1, are beginning to yield measurable benefits but have had limited cumulative impact due to their shorter implementation period and recent service disruptions. Overall, despite differences in vaccine introduction timing and occasional service disruptions, these findings underscore the critical role of routine vaccination in reducing preventable mortality in DPRK.

We presented vaccine impact estimates for 2021–2024 using two complementary approaches, applying YoV to assess the excess burdens that occurred during the pandemic and CY to evaluate the mitigating effect of catch-up vaccination. In the YoV approach, the full lifetime benefit of vaccination is assigned to the year of administration, meaning that year-by-year statistics do not reflect the actual timing of deaths and DALYs prevented, nor do they fully capture the impact of past programs. The CY approach measures outcomes by calendar year and better reflects accumulated vaccination effects. Because CY values incorporate this historical accumulation, interpreting them solely as the consequences of the 2021–2024 disruptions would risk overestimating their impact; they are more appropriately understood as reflecting the extent to which catch-up efforts partially alleviated the excess burden.

In the YoV approach, our study estimated 15 624 (95% CrI 11 379–20 747) excess deaths in DPRK during 2021–2024, compared with a 2024 multi-country analysis that estimated 967 635 (95% CrI 896 596–1 049 981) excess deaths across 112 countries for 2020–2030^3^. Despite differences in period, geographic scope, and pathogen coverage, both analyses identify interruptions in hepatitis B vaccination as the largest driver of long-term excess mortality. This illustrates that even relatively short-term declines in coverage can lead to disproportionately large health consequences that may remain hidden for years^13^, underscoring the importance of sustained primary care, catch-up vaccination, and long-term surveillance. In the CY approach, catch-up vaccination was found to reduce total excess deaths by about 1.1% and DALYs by about 0.3% in the short term. The relative reduction was largest for Pertussis (DTP1), whereas diseases with long-term vaccine effects, such as Hepatitis B and Hib, showed little immediate change. Small reductions were observed for other low-mortality diseases, but the estimates were unstable due to the low number of deaths. The mitigating effects of catch-up vaccination are expected to become more apparent over the long term, depending on each disease’s epidemiology.

Measles vaccination, historically the most impactful vaccine in the 50-year history of the Expanded Program on Immunization (EPI), has saved more lives and extended healthy life years than any other vaccine^2^, yet recent declines in coverage in DPRK, despite maintaining 92–99% coverage since the early 2000s^5^ and approaching elimination by 2018^14^, have become a major contributor to immediate and long-term health risks. According to WHO data, the reported zero measles cases in DPRK for 2022–2024 may reflect limitations in surveillance rather than the true absence of disease, highlighting the potential value of modeling to estimate health impacts^15^. In our results, CY estimates underestimate these effects due to assumptions of persistent herd immunity, whereas the YoV approach reveals the direct impact of recent coverage drops. Considering the epidemiological characteristics of measles, modeling analyses in other settings have shown that temporary disruptions in immunization can rapidly increase disease burden^16–18^. For instance, VIMC projected that postponing planned measles campaigns by one year in Ethiopia or Nigeria could markedly elevate outbreak risks, while a 50% reduction in routine coverage would further increase mortality across multiple models^19^. These findings, consistent with our YoV-based estimates, suggest that recent declines in DPRK could similarly heighten outbreak risks if not addressed through timely catch-up vaccination.

Hib3, RCV1, BCG, and DTP1 vaccines have provided steady health benefits. After hepatitis B and measles, the collapse in Hib3 coverage caused the third-largest health loss among vaccine-preventable diseases. Hib protects children under five from severe bacterial infections, including pneumonia, meningitis, and epiglottitis^20^. Introduced in 2012, coverage rose rapidly and likely reduced disease incidence^5^. Full protection requires three doses, making Hib vaccination vulnerable to disruptions during the COVID-19 pandemic, with incomplete vaccination compromising both immediate and long-term health. Hib is also a major cause of infant pneumonia, and coverage drops may raise mortality^20^. RCV1 protects reproductive-age women and infants from viral transmission and congenital rubella syndrome. While its direct contribution to excess deaths and DALYs is limited, RCV1 remains essential for infection control and child health.

BCG, given at birth, prevents severe tuberculosis in early infancy^21^. In DPRK, remained high (99%) through 2022 but declined sharply to 63% in 2023^5^. Declines could potentially increase newborn risk, although the immediate impact may be limited due to historically low TB circulation and minimal importation. DTP prevents invasive bacterial and respiratory infections in childhood, including pertussis-related deaths in infants^22^. DTP1 coverage is a key indicator of vaccine access in low-and middle-income countries^1^. While our analysis estimated relatively few excess deaths from declining DTP coverage, disease risk may have increased. Pertussis is often misdiagnosed as pneumonia, suggesting falling vaccination can produce substantial but “silent” impacts even without major outbreaks^3^. Polio vaccines were not included due to no indigenous cases since 1996, but declining coverage could increase cVDPV risk. Delivery of OPV/IPV in 2024 may help prevent this.

By estimating trends and health impacts of routine vaccination in DPRK, this study provides timely evidence to inform strategic vaccine policy. Historical patterns indicate that national crises, such as the “Arduous March” and the “COVID-19 pandemic”, have led to sharp declines in vaccine coverage. Although DPRK’s Medium Term Strategic Plan for the Health Sector (2016–2020) prioritized infection control through immunization with good quality cold chain^23^, progress was likely constrained by the exceptional circumstances of the COVID-19 pandemic^24^. In many countries, vaccination disruptions during the pandemic were attributed to health system overload or collapse. In DPRK, however, vaccination patterns may have been influenced by contextual and structural characteristics distinct from those observed elsewhere^25^.

These findings underscore the importance of maintaining or rapidly restoring routine immunization during public health emergencies. While temporary restrictions such as border closures or social distancing may help curb the immediate spread of COVID-19 and other infectious diseases, they can also increase long-term risks from vaccine-preventable diseases^26^. Recognizing both the short-term benefits and the longer-term risks of such measures is critical for balanced public health planning. Mortality often rises in socioeconomically disadvantaged areas during crises^27^, potentially widening geographic disparities. Because vaccines provide greater benefits in high-burden populations^28^, restoring coverage may be crucial for promoting health equity in DPRK.

In our study, several limitations should be considered. First, due to scarce official data from DPRK, we relied on extrapolated estimates from global sources such as WUENIC, GBD, and VIMC for coverage and incidence values, which may lead to uncertainty, particularly as GBD data may not adequately capture the immediate vaccination impacts during COVID-19, variations in health system quality, or population-specific factors. Moreover, the total estimates presented in the YoV approach should be interpreted conservatively, as they represent aggregated values derived from different data sources (VIMC and GBD). Second, the YoV and CY approaches are sensitive to assumptions about population structure, life expectancy, and disease progression. For the YoV approach, the linear relationship used may introduce more uncertainty over long-term projections. CY calculations incorporate catch-up effects through multiple probabilistic steps (e.g., CIR, CFR, vaccine efficacy), and this layered structure can generate wide variance, especially when averted deaths are few. Finally, our model did not account for contextual factors such as nutrition, COVID-19-related mobility restrictions, and healthcare facilities. In addition, it may not have fully captured the transmission dynamics of infectious diseases, nor the variation in vaccine effectiveness over time, location, and target populations, and the indirect effects resulting from reduced vaccination coverage among non-target populations (e.g., adults). However, we made efforts to consider non-target populations in the catch-up vaccination scenarios. As vaccine impact models often underestimate benefits in data-sparse or crisis-affected settings^29^, our findings should be viewed as conservative, highlighting the value of quantitative approaches for policy-making in under-researched contexts. To support interpretation and use, we followed the Guidelines for Accurate and Transparent Health Estimates Reporting (GATHER)^29^ and provided the completed checklist in Appendix G.

Moving forward, recovery strategies in DPRK must not only restore pre-pandemic coverage levels but also build systems resilient to future shocks. While vaccination declines in many countries reflected systemic collapse, in DPRK they were largely shaped by COVID-19–related border closures and restrictions on vaccine imports, which increased vulnerability to vaccine-preventable diseases. In late 2023, DPRK launched a nationwide vaccination campaign with UNICEF and Gavi support. The campaign marks important step in addressing excess mortality risks caused by prolonged restrictions. It also highlights a uniquely important case for the global immunization community, as DPRK was the only country to face such circumstances. This suggests, the importance of international collaboration and flexible support mechanisms for sustaining immunization in constrained settings.

### Research in context Evidence before this study

We searched PubMed on October 2, 2025, without date limits, using the terms “(vaccine OR immunization) AND (impact OR effect) AND model AND (mortality OR morbidity) AND COVID-19 AND (countries OR LMIC OR “low-income countries” OR “middle-income countries” OR DPRK OR DPRK)”. Studies were included if they applied modelling approaches to estimate the health impact of vaccination or pandemic-related service disruptions on vaccine-preventable diseases (VPDs) in low- and middle-income countries. Of 1,135 studies that met the inclusion criteria, five were relevant to pandemic-related immunization disruptions. One study evaluated ten vaccines across 112 LMICs using the Vaccine Impact Modelling Consortium (VIMC) framework. The other four studies focused on specific vaccines or diseases: two on HPV vaccination, one on BCG and pediatric tuberculosis, and one on dengue transmission in Southeast Asia and Latin America. None provided a systematic, pathogen-specific assessment of routine vaccination in DPRK or quantified excess burden from COVID-19–related service disruptions. While global studies reported declines in immunization, the impact at a single-country level with prolonged service interruptions remains unclear. Uncertainties persist regarding additional mortality, catch-up effectiveness, and vaccine prioritization, highlighting the need for a detailed, country-specific evaluation to inform strategic, tailored immunization policies.

### Added value of this study

This study provides the first comprehensive, pathogen-specific assessment of routine vaccination impact in DPRK over four decades (1980–2024), quantifying averted deaths and disability-adjusted life years (DALYs) for key VPDs. It also estimates the excess burden from unprecedented COVID-19–related disruptions between 2021 and 2023. Building on global analyses such as Hartner et al. (2024), which modelled disruptions across 112 countries, this study focuses in depth on a single country with severe, prolonged service interruptions, providing more granular, context-specific insights. Leveraging global modelling frameworks (VIMC and GBD) in a data-limited setting, it estimates both historical vaccine impact and pandemic-related excess burden, advancing knowledge on the impact of vaccination disruptions.

### Implications of all the available evidence

Over the past four decades, key vaccines, including hepatitis B and measles, have been particularly effective in reducing deaths and DALYs, even within a single-country context. Maintaining strategic reserves of these vaccines could provide the most effective mitigation in the event of future service disruptions. This evidence emphasizes the importance of strategically maintaining and tailoring routine immunization programs during crises, implementing effective catch-up campaigns, and strengthening health system resilience. These insights offer actionable guidance for policymakers, WHO’s Immunization Agenda 2030, and international partners such as UNICEF and Gavi to sustain immunization, reach unvaccinated populations, and prevent further excess morbidity and mortality.

## Data Availability

All data produced in the present study are available upon reasonable request to the authors
All data produced in the present work are contained in the manuscript
All data produced are available online at

## Contributors

HP conceptualised the study, developed the methods, conducted the analysis, visualization, and co-drafted the manuscript. AC reviewed the analysis, and reviewed the manuscript. YJC conceptualised the study, reviewed the analysis, and co-drafted the manuscript. SJK conceptualised the study, supported the analysis, reviewed the analysis, and co-drafted the manuscript. MC reviewed the analysis, and co-drafted the manuscript. SIB reviewed the analysis, and reviewed the manuscript. JHH reviewed the analysis, project administration, and reviewed the manuscript. YHL conceptualised the study, developed the methods, reviewed the analysis, funding acquisition, and co-drafted the manuscript. All authors had final responsibility to submit the manuscript for publication.

## We declare no competing interests

### Data sharing

All models were calibrated with publicly available datasets as described in the article and references for each model.

## Acknowledgments

This work was supported by grants from the National Research Foundation of Korea (NRF) under the Ministry of Science and Information and Communication Technology [grant number RS-2023-00249082] and Korea University. We are grateful to Professor Kaja Abbas (London School of Hygiene & Tropical Medicine) for his careful review of the manuscript.

## Notes

### Competing Interest Statement

The corresponding author is supported by the National Research Foundation of Korea and Korea University.

